# Optimal duration of antibiotic treatment for community-acquired pneumonia in adults: a systematic review and duration-effect meta-analysis

**DOI:** 10.1101/2021.12.09.21267441

**Authors:** Yuki Furukawa, Yan Luo, Satoshi Funada, Akira Onishi, Edoardo G Ostinelli, Tasnim Hamza, Toshi A Furukawa, Yuki Kataoka

**Author notes:** **Corresponding author** Dr. Yuki Furukawa, MD., Tokyo Musashino Hospital, 4-11-11, Komone, Itabashi-ku, Tokyo, 173-0037, Japan.

## Abstract

**Importance:** Community-acquired pneumonia (CAP) is a leading cause of morbidity and mortality globally. The optimal duration of antimicrobial therapy remains unclear and controversial.

**Objective:** To find the optimal treatment duration with antibiotics for CAP in adults.

**Data Sources:** MEDLINE, Embase and CENTRAL from inception to present (25 August, 2021).

**Study Selection:** All randomized controlled trials comparing the same antibiotics used at the same daily dosage but for different durations for CAP in adults. We included any antibiotics, administered orally or intravenously. We included both outpatients and inpatients but not those admitted to intensive care unit.

**Data Extraction and Synthesis:** Two review authors independently screened and extracted data. We conducted random-effects, one-stage duration-effect meta-analysis with restricted cubic splines. We tested the non-inferiority with the pre-specified non-inferiority margin of 10% examined against 10 days using intention-to-treat dataset.

**Main Outcomes and Measures:** The primary outcome was clinical improvement at day 15 (range 7-45 days). Secondary outcomes were all-cause mortality, serious adverse events, and clinical improvement at day 30 (15-60 days). We calculated odds ratios.

**Results:** We included 9 trials (2399 patients with a mean [SD] age of 61.2 [22.1]; 39% women). The duration-effect curve was monotonic with longer duration leading to lower probability of improvement, and the lower 95%CI curve was constantly above the prespecified non-inferiority margin throughout the examined duration. Harmful outcome curves indicated no association. The average percentage of clinical improvement rate at day 15 in the 10-day treatment arms was 68%. Using that average, we computed the absolute clinical improvement rates at the following durations: a 3-day treatment 75% (95%CI: 68 to 81%), 5-day treatment 72% (66 to 78%), and 7-day treatment 69% (61 to 76%).

**Conclusions and Relevance:** Shorter treatment duration probably achieves the optimal balance between efficacy and treatment burden for treating CAP in adults. However, the small number of included studies and the overall moderate to high risk of bias may compromise the certainty of the results. Further research focusing on the shorter duration range is required.

**Registration:** PROSPERO (CRD 42021273357).

**KEY POINTS:** *Question:* What is the optimal treatment duration of antibiotics for community-acquired pneumonia (CAP) in adults.?

*Findings:* This systematic review and duration-effect meta-analysis of 9 trials with 2399 patients found that the shorter treatment duration (3-9 days) was likely to be non-inferior to the standard treatment duration (10 days) for CAP in adults if they achieved clinical stability.

*Meaning:* Shorter antibiotic treatment duration probably achieves the optimal balance between efficacy and treatment burden for CAP in adults.

## INTRODUCTION

Community-acquired pneumonia (CAP) is a leading cause of morbidity and mortality globally, especially among the elderly.^1^ In the United States, it is the second most common cause of hospitalization and the top infectious cause of death.^2,3^ The initial treatment for CAP is empirical, with guidelines recommending starting several antibiotics depending on patients’ severity and risk factors for certain pathogens.^4–6^

The optimal duration of antimicrobial therapy remains unclear and controversial. The American and British guidelines recommend a minimum of five days of treatment before therapy discontinuation for patients achieving clinical stability.^4,5^ The European guideline states that the duration of treatment should not exceed 8 days in responding patients.^6^ In clinical practice, however, antibiotics for pneumonia are often prescribed for 10 up to 14 days.^7,8^ This may mean that many patients may be receiving more antibiotics than necessary, with a consequent increase in costs and a higher probability of antimicrobial resistance.^9^ Finding optimal duration of antibiotics can facilitate reducing antimicrobial use efficiently. A pair-wise meta-analysis published in 2008 found that short-course therapy was non-inferior to long-course therapy regarding clinical success at end-of-therapy, clinical success at late follow-up, microbiological success, relapses, mortality and adverse events.^10^ Since then, at least two trials have been reported,^11,12^ which warrants update of the systematic review and meta-analysis. A major limitation of the method used in the previous pair-wise meta-analysis is the arbitrary categorization of durations when the original studies compared different durations, ranging from three to ten days. This resulted in categorizing a seven-day treatment in one trial to short-course and the same in other two trials to long-course.^13–15^

We overcame this limitation by using a novel method called dose-effect meta-analysis.^16^ It has been used, for example, to identify optimal dose of antidepressants for major depressive disorder^17^ and antipsychotics for schizophrenia^18^. Unlike conventional categorization-based meta-analyses^19^, dose-effect meta-analysis can reveal more fine-grained optimal dose^20^. By treating duration as dose, we aimed to apply this method to obtain a more specific optimal treatment duration.

## OBJECTIVE

To find the optimal treatment duration of antibiotics for CAP in adults.

## METHODS AND ANALYSIS

We followed the Preferred Reporting Items for Systematic reviews and Meta-Analyses (PRISMA 2020) ^21^. The protocol has been prospectively registered in PROSPERO (CRD 42021273357) and can be found in the appendix (eAppendix1).

### Data sources

#### Criteria for considering studies for this review

##### Types of studies

To examine the duration-effect relationship, we included all trials that compared two or more different durations of the same antibiotic treatment for CAP.

##### Types of participants

Patients were eligible if they were 18 years or older of both genders with a diagnosis of CAP as defined by the original authors. We included both outpatients and inpatients. We excluded patients who were admitted to intensive care unit. In order to focus on individuals at low to medium risk, we excluded trials with 20% or more patients meeting one or more of the following criteria: having immunodeficiency; having been treated with another antibiotic within a month.

##### Types of interventions

We included trials examining any antibiotics, administered orally or intravenously. We evaluated antibiotics as a class because clinical guidelines recommend treatment duration irrespective of the antibiotic used,^4–6^ and because recent meta-analyses of antibiotics for CAP have not shown efficacy differences among antibiotics.^22,23^ Oral and intravenous antibiotics were merged because they have been shown equally effective in many infectious conditions within the same time frame.^24–26^ We included trials comparing the same agents used at the same daily dosage but for different durations. We used the predefined duration for fixed-duration arms. If some studies did not prespecified the duration (eg. left it to clinicians’ judgment^11^), we used the median duration.

### Primary outcome and secondary outcomes

The primary outcome of interest in this study was clinical improvement as defined by the original authors at a time point as close to 15 days (range 7-45 days) as possible in each included study.^27^ Secondary outcomes of interest were: all-cause mortality at day 15 (range 7-45 days), serious adverse events as defined by the original study at day 15 (range 7-45 days), and clinical improvement as defined by the original study at day 30 (range 15-60). We used the number of randomized patients as the denominator for intention-to-treat (ITT) dataset. When only clinical failure was reported, clinical improvement was calculated by subtracting clinical failure from the total number randomized. We used ITT for the primary analysis and per-protocol (PP) dataset for a sensitivity analysis.^28,29^ We used odds ratio (OR) of each outcome to synthesize data.^30,31^

#### Search methods for identification of studies

##### Electronic searches

We systematically searched the following electronic bibliographic databases from inception to present (25 August, 2021): MEDLINE, Embase and CENTRAL. We used search terms for community acquired pneumonia in conjunction with the names of individual antibiotics as well as the names of antibiotic classes. Detailed search formulas are presented in the appendix (eAppendix2). We imposed no date, language or publication status restriction.

##### Reference lists

We checked the reference lists of all the included studies and review articles for additional references.

### Data collection and analysis Selection of studies

Two review authors independently screened and selected the included studies (YF and one of AO, EO, SF or YL). Two review authors extracted data independently from the included studies (YF and one of AO, EO, SF or YL). We used the Cochrane risk of bias tool Version 2 ^32^ to assess and summarize the risk of bias. Disagreements were resolved through discussion.

### Statistical analysis

To perform our analyses, we used the *dosresmeta* package (Version 2.0.1) and *meta* package (Version 5.0-1) for *R* (Version 4.1.0. R foundation, Wien, Austria).^33–35^

#### Assessment of heterogeneity

We investigated the heterogeneity between studies by the variance partition coefficient (VPC). ^16^ VPC represents the percentage of variation attributed to heterogeneity rather than sampling error and can be interpreted similarly to the I^2^.

#### Dose-effect meta-analysis

Given the clinical and methodological heterogeneity likely present in the included studies, we used the random effects model. We used 3 knots, equally spaced across the duration range (25%, 50%, 75%).^17,18^ We set 10 days as the reference because it can be regarded as the current practice.^7,8,11^ We tested the non-inferiority with the non-inferiority margin of 10%, as previously proposed,^27^ and the superiority of the shorter duration examined against 10 days using ITT dataset.

#### Sensitivity analyses

In order to ascertain the robustness of the primary analyses, we conducted the following sensitivity analyses. To test the stability of the shape of the spline curves, we used different locations of knots (10%, 50%, 90%). To test the influence of trials included, we conducted sensitivity analyses excluding trials with an overall high risk of bias and excluding trials with outpatients. To test the robustness of the analytical method, we used the PP dataset. To test the influence of antibiotics examined, we conducted sensitivity analyses restricting eligible antibiotics only to those recommended by clinical guidelines for empirical treatment of CAP.^4,5^ In addition to the pre-defined sensitivity analyses, we conducted exploratory sensitivity analyses including only trials that randomized before the initial antibiotic treatment to test the influence of randomization timing.

### Amendments

We report amendments with the date and the rationale in the appendix (eAppendix3).

## RESULTS

We identified 1,994 records via database and one record via searching websites, which revealed that some different records refer to the same clinical trial. We assessed 38 full-text records for eligibility and included 11 eligible studies. (Fig1) Of these, 8 were published,^11–15,36–38^ 1 was unpublished^39^ and 2 studies were still ongoing,^40,41^ resulting in 9 trials for the primary outcome analysis. The lists of included and excluded studies are provided in the appendix (eAppendix4 and 5). The 9 studies with 2,399 participants in total included 18 eligible arms. Treatment duration ranged from 3 to 10 days. The study year ranged between 1999 and 2021. Table 1 presents the characteristics of the included studies.

**Figure 1.**
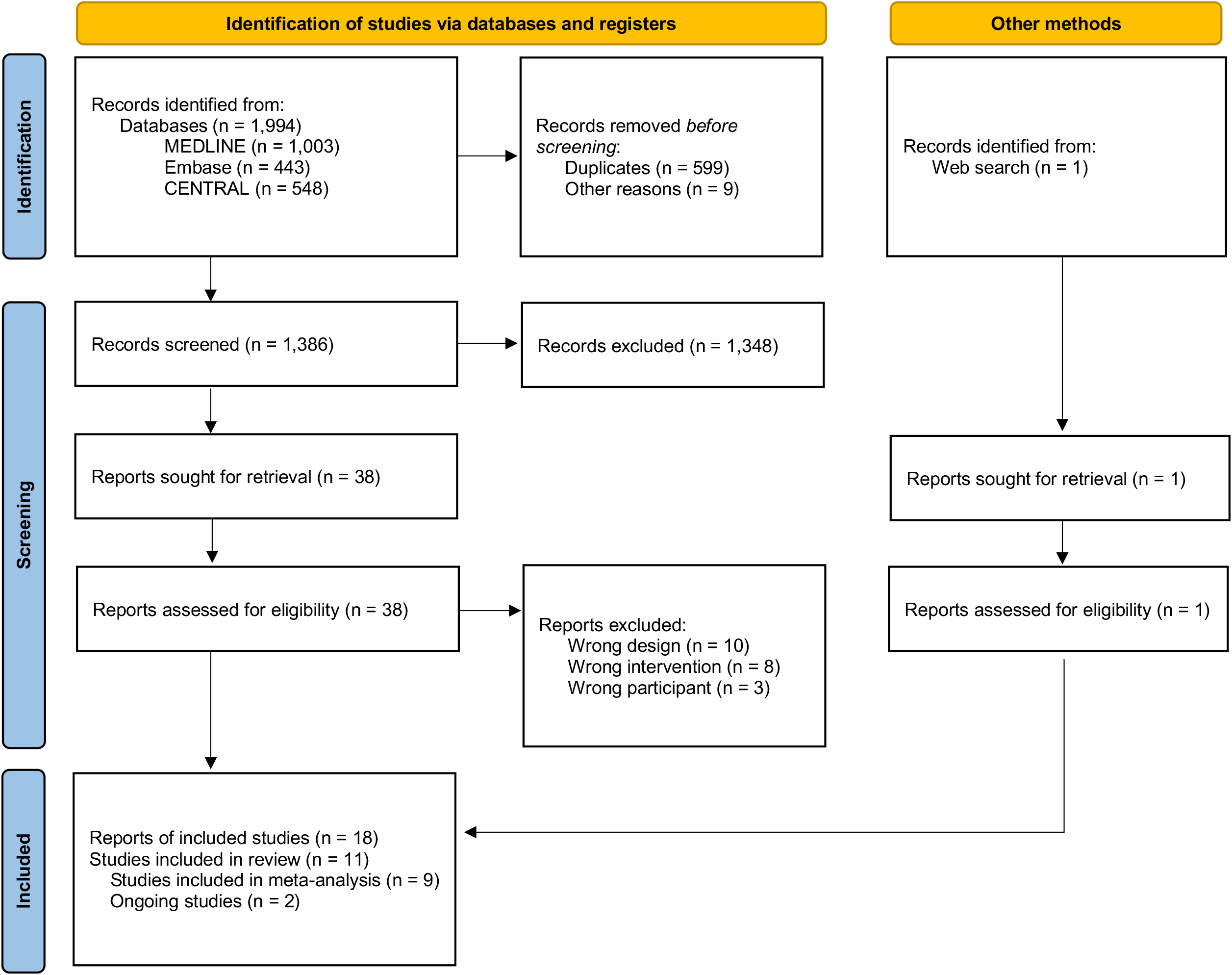
PRISMA flow diagram.

**Table 1.**
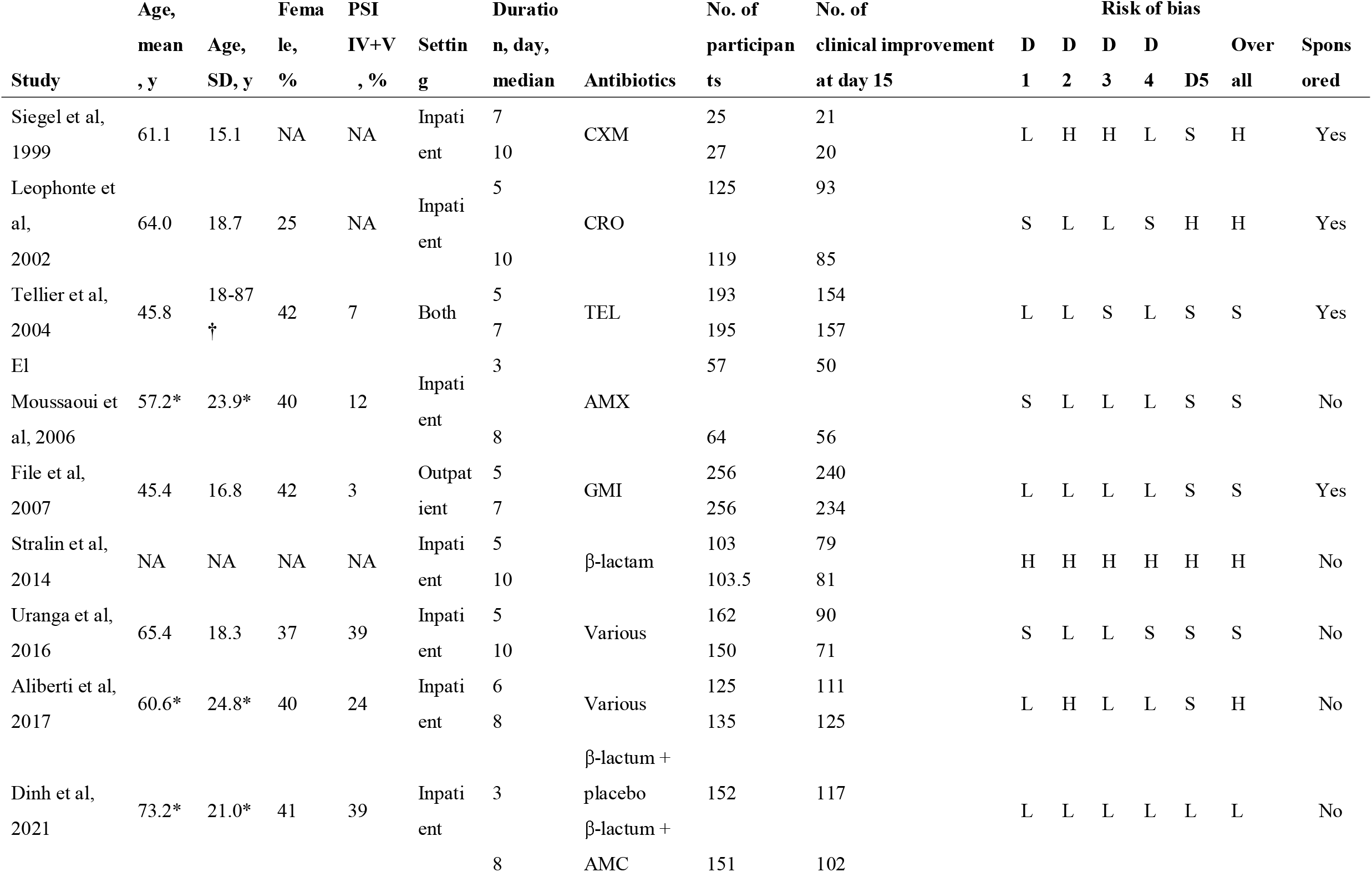
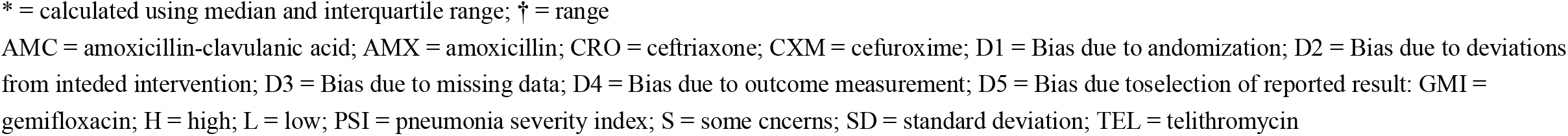
Characteristics of included studies.

| Study | Age, |  | Fema<br>le,<br>% | PSI<br>IV+V<br>, % | Settin<br>g | Duratio<br>n, day,<br>median | Antibiotics | No. of<br>participan<br>ts | No. of<br>clinical improvement<br>at day 15 | Risk of bias |  |  |  |  | Over<br>all | Spons<br>ored |
| --- | --- | --- | --- | --- | --- | --- | --- | --- | --- | --- | --- | --- | --- | --- | --- | --- |
|  | mean<br>, y | Age,<br>SD, y |  |  |  |  |  |  |  | D<br>1 | D<br>2 | D<br>3 | D<br>4 | D5 |  |  |
| Siegel et al,<br>1999 | 61.1 | 15.1 | NA | NA | Inpati<br>ent | 7<br>10 | CXM | 25<br>27 | 21<br>20 | L | H | H | L | S | H | Yes |
| Leophonte et<br>al,<br>2002 | 64.0 | 18.7 | 25 | NA | Inpati<br>ent | 5<br>10 | CRO | 125<br>119 | 93<br>85 | S | L | L | S | H | H | Yes |
| Tellier et al,<br>2004 | 45.8 | 18-87<br>† | 42 | 7 | Both | 5<br>7 | TEL | 193<br>195 | 154<br>157 | L | L | S | L | S | S | Yes |
| El<br>Moussaoui et<br>al, 2006 | 57.2* | 23.9* | 40 | 12 | Inpati<br>ent | 3<br>8 | AMX | 57<br>64 | 50<br>56 | S | L | L | L | S | S | No |
| File et al,<br>2007 | 45.4 | 16.8 | 42 | 3 | Outpat<br>ient | 5<br>7 | GMI | 256<br>256 | 240<br>234 | L | L | L | L | S | S | Yes |
| Stralin et al,<br>2014 | NA | NA | NA | NA | Inpati<br>ent | 5<br>10 | β-lactam | 103<br>103.5 | 79<br>81 | H | H | H | H | H | H | No |
| Uranga et al,<br>2016 | 65.4 | 18.3 | 37 | 39 | Inpati<br>ent | 5<br>10 | Various | 162<br>150 | 90<br>71 | S | L | L | S | S | S | No |
| Aliberti et al,<br>2017 | 60.6* | 24.8* | 40 | 24 | Inpati<br>ent | 6<br>8 | Various | 125<br>135 | 111<br>125 | L | H | L | L | S | H | No |
| Dinh et al,<br>2021 | 73.2* | 21.0* | 41 | 39 | Inpati<br>ent | 3 | β-lactum +<br>placebo | 152 | 117 | L | L | L | L | L | L | No |
|  |  |  |  |  |  | 8 | β-lactum +<br>AMC | 151 | 102 |  |  |  |  |  |  |  |

\* = calculated using median and interquartile range; † = range AMC = amoxicillin-clavulanic acid; AMX = amoxicillin; CRO = ceftriaxone; CXM = cefuroxime; D1 = Bias due to andomization; D2 = Bias due to deviations from inteded intervention; D3 = Bias due to missing data; D4 = Bias due to outcome measurement; D5 = Bias due toselection of reported result: GMI = 0 gemifloxacin; H = high; L = low; PSI = pneumonia severity index; S = some cncerns; SD = standard deviation; TEL = telithromycin

The included studies were all parallel-group and individually randomized. Seven out of nine were reported as non-inferiority trial. In total, 1,199 participants were randomly assigned to the shorter duration arm and 1,200 to the longer duration arm. The mean age was 61.2 years (standard deviation 22.1); 831 (39%) of 2,140 reported were women. Six were conducted in a single European country, one in the US, and the two were cross-continental. CAP was defined as newly confirmed clinical symptoms (eg, dyspnoea, cough, purulent sputum, or crackles), and radiological findings. Clinical stability was often defined as apyrexia (temperature ≤37.8□C) for 48 hours, heart rate below 100 beats per min, respiratory rate below 24 breaths per min, arterial oxygen saturation of 90% or higher, systolic blood pressure of 90 mm Hg or higher, and normal mental status.^42^ Percentage of pneumonia severity index class IV or V was on average 19% (362 of 1,896 reported; ranging from 2 to 41%). Seven studies focused on inpatients, whereas one study focused on outpatients and one included both. Antibiotics used included β-lactam (amoxicillin, amoxicillin/clavulanate, ampicillin/sulbactam, ceftazidime, ceftriaxone, cefuroxime, piperacillin/tazobactam), macrolide (azithromycin, clarithromycin), quinolone (ciprofloxacin, gemifloxacin, levofloxacin, telithromycin), amikacin, doxycycline, and meropenem. Pharmaceutical companies funded four studies.^13–15,36^ Four studies had a high overall risk of bias, four some concerns, and only one had low overall risk of bias. (Table 1)

### Assessment of heterogeneity

We assessed the heterogeneity in efficacy outcome across duration range (9 studies). VPC values were constantly below 10% which suggests low levels of heterogeneity. However, these assessments need to be carefully interpreted due to the small number of included studies. (eAppendix6)

### Dose-effect meta-analysis

We present the duration-effect curves in Figure 2 and 3, and the tabulation of results in Table 2. The x-axis of the figures represents the treatment duration in days and the y-axis represents the odds ratio of the outcome. The thin solid horizontal line represents the odds ratio = 1 and the thin dotted horizontal line in the clinical improvement figures corresponds to the non-inferiority margin translated into OR. (The average percentage of clinical improvement rate at day 15 in the 10-day treatment arms was 68%. Non-inferiority margin was therefore 58% and the corresponding OR was 0.65. For clinical improvement at day 30, the numbers were 77%, 67% and OR 0.61, respectively.) The thick solid line represents the dose-effect curve and the thick dotted lines represent its 95% CI. The duration-effect curve is monotonic with longer duration leading to lower probability of improvement. The lower 95%CI curve was constantly above the prespecified non-inferiority margin, meaning that a shorter treatment duration (3-9 days) was likely to be non-inferior to the standard treatment duration (10 days). It was slightly above the OR = 1 line around 3 days, suggesting 3-day treatment may be superior to 10-day treatment. Secondary outcomes had wider confidence interval curves. Harmful outcome curves (all-cause mortality and severe adverse events) were almost flat and 95%CI curves did not cross the OR = 1 line, indicating no association. Clinical improvement at day 30 showed a similar trend with the primary outcome with the lower 95%CI curve constantly above the prespecified non-inferiority margin. The average percentage of clinical improvement rate at day 15 in the 10-day treatment arms was 68% (based on a meta-analysis of the included studies). Using that average, we computed the absolute clinical improvement rates at the following durations: a 3-day treatment 75% (95%CI: 68 to 81%), 5-day treatment 72% (66 to 78%), and 7-day treatment 69% (61 to 76%).

**Table 2.**
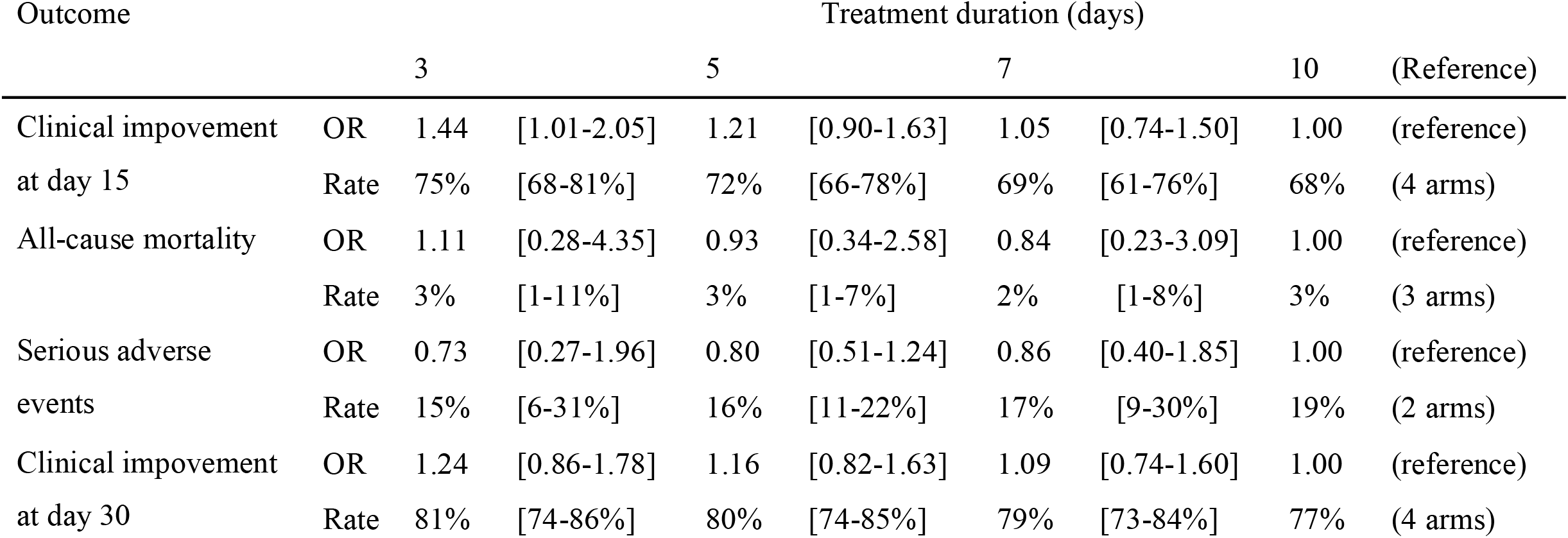
Tabulation of results.

**Figure 2.**
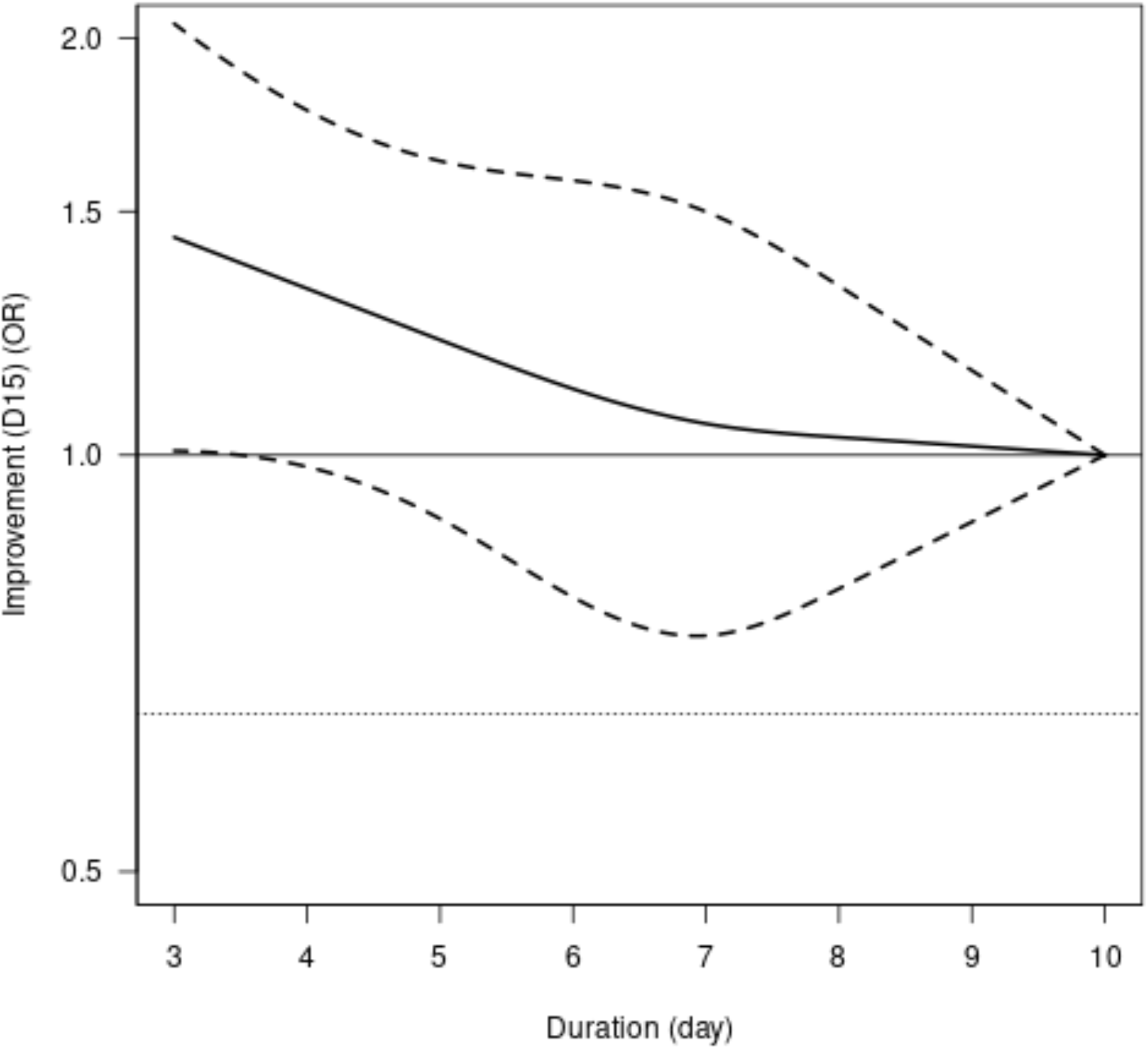
Duration–effect relationship of antibiotics for CAP in adults. Clinical improvement at day 15. OR=odds ratio. The dotted lines represent 95% confidence intervals. The thin horizontal dotted line represents the non-inferiority margin, corresponding with 10% absolute risk difference given the average event rate of 68%

**Figure 3.**
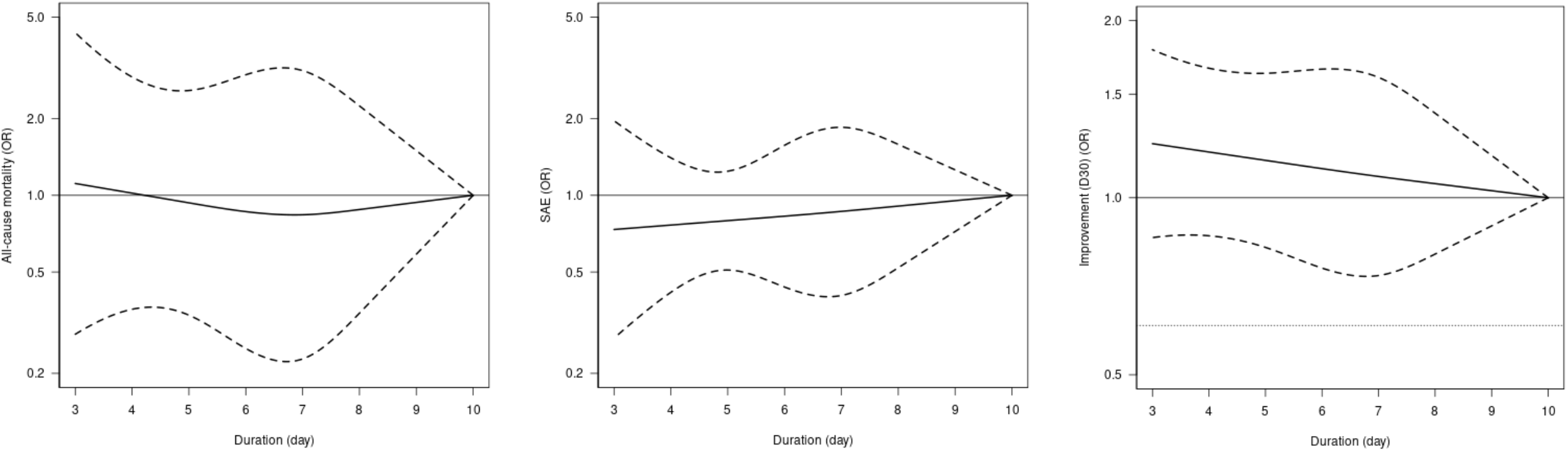
Duration–effect relationships of antibiotics for CAP in adults. (a) All-cause mortality. (b) Severe adverse events. (c) Clinical improvement at day 30. OR=odds ratio. The dotted lines represent 95% confidence intervals. The thin horizontal dotted line represents the non-inferiority margin, corresponding with 10% absolute risk difference given the average event rate of 77%

### Sensitivity analyses

Sensitivity analyses were in line with the primary analyses. (eAppendix7. Figures S1, using different locations of knots; S2.1, excluding trials with overall high risk of bias; S2.2, excluding trials with outpatients; S3, using PP dataset; S4 including only antibiotics recommended for empirical treatment of CAP by clinical guidelines). Exploratory sensitivity analyses showed that non-inferiority of the shorter duration was more likely to be the case in studies that randomized patients who had reached clinical stability early (eAppendix7. Figures S5.1, S5.2).

## DISCUSSION

To our knowledge, this is the first systematic review and duration-effect meta-analysis of antibiotics treatment for CAP in adults. The results showed that a shorter treatment duration (3-9 days) was likely to be non-inferior to the standard treatment duration (10 days) for CAP in adults. There may be no significant difference in all-cause mortality or serious adverse events. A shorter range probably achieves the optimal balance between efficacy and treatment burden.

This is in line with the previous pair-wise meta-analysis that showed shorter duration was non-inferior to longer duration.^10^ Methodological limitations in a previous meta-analysis restricted authors from recommending a specific treatment duration. We overcame this limitation by examining the duration of antibiotic treatment range in days and found that a 3 to 9-day treatment is likely to be non-inferior to a 10-day treatment. Our results are in line with the guidelines for CAP recommending antibiotics to be prescribed for a duration shorter (5-8 days) than current clinical standard practice (10 days).^4–6^ Our results suggest that an even shorter duration (3-5 days) may be considered, which is in line with the trials that found 3-day treatment was non-inferior to 8-day treatment.^12,37^

### Limitations

Our study has several limitations. First, most of the included studies presented with moderate to high overall risk of bias. Second, the number of studies was small, leaving confidence intervals for secondary outcomes wide. Third, original studies excluded patients with complications of CAP and therefore the results of this study may not be generalizable to those patients. Forth, baseline severity of the included studies varied. However, the overall heterogeneity was low.

### Strengths

First, we did a comprehensive systematic review and found 4 studies that were not included the previous systematic reviews. Second, we treated duration as a continuous variable, which allowed us to estimate the duration-effect relationship with greater resolution of change points. Third, we examined impacts of treatment duration not only for clinical improvement but also for all-cause mortality and severe adverse events and made sure that a shorter treatment duration would not translate into more harmful events. Finally, the very nature of shortened duration treatment offers a unique opportunity for interpretation. Shorter treatment durations have been examined by non-inferiority trials. The underlying assumption has been that there was a trade-off between a loss in efficacy of standard treatment duration and other benefits of a shortened duration, ^43,44^ such as less time, less cost and probably a diminished rate of antimicrobial resistance. This study suggests that there may be even no trade-off for antibiotic treatments of 3 to 5 days. Shorter treatment duration reduces the burden on patients, the healthcare system and the risk of antimicrobial resistance and might even offer better clinical outcomes at the same time.

## CONCLUSION

Shorter treatment duration (3-9 days) was likely to be non-inferior to the standard treatment duration (10 days) for adults with CAP if they achieved clinical stability. A shorter range (3-5 days) probably results in an optimal balance between efficacy and treatment burden. However, the small number of included studies and the overall moderate to high risk of bias may compromise the certainty of the results. Further research focusing on the shorter duration range is required.

## Supporting information

eAppendix

## Data Availability

All data produced in the present study are available upon reasonable request to the authors

## Abbreviations

CAP: community-acquired pneumonia
ITT: intention-to-treat
PP: per protocol
PRISMA: Preferred Reporting Items for Systematic Reviews and Meta-analyses
VPC: variance partition coefficient

## Registration

This protocol was prospectively registered in PROSPERO (CRD 42021273357).

## Patient and public involvement

Patients or the public were not involved in the design, conduct, reporting or dissemination plans of this research.

## Ethics

This study uses published aggregate data and did not require ethical approval.

## CRediT contribution statement

**Yuki Furukawa**: Conceptualization, Methodology, Software, Validation, Formal analysis, Data Curation, Writing – Original Draft, Visualization, Supervision, Project administration

**Yan Luo**: Conceptualization, Methodology, Data Curation, Validation, Writing – Review & Editing,

**Satoshi Funada**: Conceptualization, Methodology, Data Curation, Validation, Writing – Review & Editing,

**Akira Onishi**: Conceptualization, Methodology, Data Curation, Validation, Writing – Review & Editing,

**Edoardo G Ostinelli**: Conceptualization, Methodology, Data Curation, Validation, Writing – Review & Editing,

**Tasnim Hamza**: Methodology, Software, Formal analysis, Visualization, Validation, Writing – Review & Editing,

**Toshi A Furukawa**: Conceptualization, Methodology, Writing – Review & Editing, Supervision,

**Yuki Kataoka**: Conceptualization, Methodology, Writing – Review & Editing, Supervision,

## Data availability

Data and code used for analyses are available from the corresponding author on reasonable request.

