## Supplementary material for "Optimal duration of antibiotic treatment for community-acquired pneumonia in adults: a systematic review and duration-effect meta-analysis": eAppendix

2. Search strings used for Ovid MEDLINE, Embase, and CENTRAL.

3. Amendments from the protocol

4. List of all included papers

5. List of excluded studies

6. Heterogeneity: Variance partition coefficient for the primary outcome

7. Sensitivity analyses

**1. Optimal duration of antibiotic treatment for community-acquired pneumonia in adults: protocol for a systematic review and duration-effect network meta-analysis (protocol as of 15^th^ August, 2021)**

Yuki Furukawa, Yan Luo, Satoshi Funada, Akira Onishi, Edoardo G Ostinelli, Tasnim Hamza, Toshi A Furukawa, Yuki Kataoka

**INTRODUCTION**

Community-acquired pneumonia (CAP) continues to be a leading cause of morbidity and mortality globally. (1) In the United States, for example, it is the second most common cause of hospitalization and the top infectious cause of death. (2,3) Clinical guidelines recommend starting several antibiotics empirically for non-severe pneumonia. (4) The optimal duration of antimicrobial therapy, however, remains unclear and controversial. Recent clinical guidelines suggest a minimum of five days of treatment before therapy discontinuation for patients achieving an afebrile state for 48 to 72 hours and meeting clinical stability criteria. (4) In clinical settings, however, a conventional ten to 14-day therapy is still used. (5,6) This may mean that many patients are receiving more antibiotics than necessary, which leads to an increased cost, time and also, higher probability of antimicrobial resistance. (7) Finding optimal duration of antibiotics is therefore meaningful not only for clinicians but also for policy-makers. A meta-analysis found that short-course therapy was not inferior to long-course therapy. (8) A major limitation of the method used in this meta-analysis is the arbitrary categorization of durations, when the original studies compared different durations, ranging from three to ten days. This resulted in categorizing a seven-day treatment in one trial to short-course and the same in another trial to long-course. We can overcome this limitation by using a novel method called dose-effect network meta-analysis (DE-NMA), which allows us to use the original duration in days and to examine the optimal duration with greater resolution of change points.

**OBJECTIVES**

To find the optimal treatment duration with antibiotics for CAP.

**METHODS AND ANALYSIS**

We follow PRISMA-P in reporting the protocol and will follow PRISMA(9) and PRISMA-NMA in reporting the DE-NMA results.

**Data sources**

**Criteria for considering studies for this review**

***Types of studies***

All randomized controlled studies. Quasi-randomized trials (such as those allocating by using alternate days of the week) will be excluded.

1. Cluster-randomized trials

Cluster-randomized trials will be included as long as proper adjustment for the intra-cluster correlation is conducted in accordance with the Cochrane Handbook for Systematic Reviews of Interventions.

2. Studies with multiple treatment groups

Where multiple trial arms are reported in a single trial, we will include only the relevant arms.

***Types of participants***

Patients of 18 years or older of both sexes with diagnosis of CAP as defined by the original authors. We will include both outpatients and inpatients. We will exclude patients who are admitted to intensive care unit. In order to focus on population without an elevated risk, we will exclude trials with 20% or more patients meeting one or more of the following criteria: having immunodeficiency; having been treated with another antibiotic within a month.

***Types of interventions***

We will include trials examining any of the antibiotics, administered orally or intravenously. As we can expect a limited number of studies to include, we will not be able to evaluate individual antibiotics. We will evaluate antibiotics as a class because clinical guidelines recommend treatment duration irrespective of the antibiotic used, (4) and because recent meta-analyses of antibiotics for CAP have not shown efficacy difference among antibiotics. (10,11) Oral and intravenous antibiotics will be merged, because they have been shown equally effective in many infectious conditions. (12–15) We will include trials comparing the same agents used in the same daily dosage but for different durations. We will use the predefined duration for fixed-duration arms and median duration for flexible-duration arms. If median duration is not reported, we will use mean duration. We will prioritize median duration because patients requiring longer duration may inflate the mean duration in flexible-duration arms.

**Primary outcome and secondary outcomes**

The primary outcome of interest in this study is clinical improvement as defined by the original authors at a time point as close to 15 days (range 7-45 days) as possible in each included study. (16) If equidistant, we will use the longer timeframe.

1 Clinical improvement at day 15 (range 7-45 days), as defined by the original study

Secondary outcomes of interest are the following outcomes.

2. All-cause mortality at day 15 (range 7-45 days)

3. Serious adverse events as defined by the original study at day 15 (range 7-45 days)

4. Clinical improvement, as defined by the original study, at day 30 (range 15-60)

We will use the number of randomized patients as the denominator for intention-to-treat (ITT) dataset and we will use per-protocol (PP) dataset as defined by the original study. Those who had been randomized but not accounted for in the original study will be assumed to have dropped out for some reason other than death or serious adverse events and without clinical improvement. In case only one of PP or ITT can be obtained, we will use the same number for the other. We will use ITT for the primary analysis and PP for a sensitivity analysis. (17,18)

**Search methods for identification of studies**

***Electronic searches***

Searches for published studies will be undertaken in the following electronic bibliographic databases from inception to present (25 August, 2021): Ovid MEDLINE and Cochrane CENTRAL. We will use search terms for community acquired pneumonia in conjunction with the names of individual antibiotics as well as the names of antibiotic classes. We imposed no date, language or publication status restriction.

***Search formula***

Search strategy for Ovid MEDLINE is as follows

#1 randomized controlled trial.pt.

#2 controlled clinical trial.pt.

#3 randomized.ab.

#4 placebo.ab.

#5 drug therapy.fs.

#6 randomly.ab.

#7 trial.ab.

#8 groups.ab.

#9 or/#1-#8

#10 exp animals/ not humans.sh.

#11 #9 not #10

#12 exp Community-Acquired Infections/

#13 Pneumonia, Bacterial/dt [Drug Therapy]

#14 community acquired pneumonia.ab,ti.

#15 (#12 and #13) or #14

#16 ((short adj term) or (long adj term) or prolonged or (short adj course) or (long adj course) or day or days or duration or disconti*).mp.

#17 (beta-lactam* or macrolide* or quinolone* or tetracycline* or amikacin or amoxicillin or ampicillin or azithromycin or cefepim or cefotaxim* or ceftarolin or ceftazidim* or ceftibuten or ceftriaxon* or cefuroxim* or cethromycin or ciprofloxacin or clarithromycin or clavulanic acid or clindamycin or co-amoxiclav or co-trimoxacol or doxycyclin* or ertapenem or erythromycin or fluoroquinolon* or fluorchinolon* or gemifloxacin or gentamicin or imipenem or levofloxacin or linezolide or meropenem or moxifloxacin or penicillin* or piperacillin or roxithromycin or sultamicillin or tazobactam or telithromycin or tetracyclin* or ticarcillin or tobramycin).mp.

#18 Anti-Bacterial Agents/ad [Administration & Dosage]

#19 #17 or #18

#20 #11 and #15 and #16 and #19

***Reference lists and others***

We will check the reference lists of all the included studies and review articles for additional references. We will also contact experts in the field to identify unpublished and on-going trials.

**Data collection and analysis**

**Selection of studies**

Two review authors will independently screen titles and abstracts of all the potential studies we identify as a result of the search and code them as 'retrieve' (eligible or potentially eligible/unclear) or 'do not retrieve'. We will retrieve the full text study reports/publication and two review authors will independently screen the full text and identify studies for inclusion and identify and record reasons for exclusion of the ineligible studies. We will resolve any disagreement through discussion or, if required, through consultation with a third review author. We will identify and exclude duplicates of the same study so that each study rather than each report is the unit of analysis in the review. We will record the selection process in sufficient detail to complete a PRISMA flow diagram and characteristics of excluded studies table.

**Data items**

We will use a standardized data collection form for study characteristics and outcome data which will have been piloted on at least one study in the review. Two review authors will extract data independently from the included studies. Any disagreement will be resolved through discussion, or discussed with a third person if necessary. We will abstract the following information.

1. ***Characteristics of the studies***

Name of the study, year of publication, country, study site (single or multi-center), study design, patient characteristics (mean age, percentage of women, diagnostic criteria used), outcome (definition of clinical success), definition of clinical stability, timing of randomization, sponsorship (rated positive if the trial is directly sponsored by drug company or if any authors are employed by the drug company).

2. ***Risk of bias***

We will use Cochrane Risk of Bias 2.0 tool (RoB2) (19). We will assess the effect of assignment to the interventions at baseline because we use the ITT population in our primary analysis.

3. ***Data to calculate effect sizes and conduct dose-effect network meta-analysis***

Patients (number of participants randomized to each arm)

Interventions (placebo or name and the dose and duration of the drug used)

Outcomes (number of clinical success, mortality, adverse events).

**Statistical analysis**

***Assessment of the network transitivity, consistency, heterogeneity and publication bias***

We will evaluate

1) transitivity of the network by comparing potential effect modifiers (severity, comorbidity, age) across comparisons

2) consistency by global as well as local tests of inconsistency

3) heterogeneity by common tau

We decided not to draw a funnel plot, because there is no appropriate method to draw it in DE-NMA and even if there is, it would be uninterpretable.

***Dose-effect network meta-analysis***

We will then conduct a DE-NMA with the *MBNMAdose* package in R.(20,21) One advantage of the dose-effect network meta-analysis by *MBNMAdose* package is that we can connect nodes that might otherwise be disconnected, by linking up different durations via the duration-effect relationship.(20) Given the clinical and methodological heterogeneity likely present in the included studies, we will use the random effects model. We will use 3 knots, equally spaced across the duration range (25%, 50%, 75%), because we do not know a priori where the outcomes change. We will test different knot placements in sensitivity analyses. We will use odds ratio of each outcome to synthesize data. (22,23)

We will set 10 days as the reference, because it is the current practice. (5,6,24) We will test the non-inferiority of the shorter duration examined against 10 days using ITT dataset, with the non-inferiority margin of 10%, as previously proposed. (16) We will compare the margin and the 95% confidence interval. In case non-inferiority is shown, we will test the superiority of the shorter duration examined against 10 days.

***Sensitivity analyses***

In order to ascertain the robustness of the primary analyses, we will conduct the following sensitivity analysis and subgroup analysis.

1 To test the stability of the shape of the spline curves, using different numbers and locations of knots

2 To test the influence of trials included,

2.1 excluding trials with overall high risk of bias

2.2 excluding trials with inpatients

3 To test the robustness of the analytical method, using PP dataset

4 To test the influence of antibiotics examined, including only antibiotics recommended for empirical treatment of CAP by clinical guidelines: beta-lactam (amoxicillin, amoxicillin/clavulanate ampicillin/sulbactam, cefotaxime, ceftriaxone, ceftraroline), macrolide (azithromycin , clarithromycin), doxycycline, respiratory fluoroquinolone (levofloxacin, moxifloxacin, gemifloxacin)

**Patient and public involvement**

Patients or the public were not involved in the design, conduct, reporting or dissemination plans of this research.

**Ethics and dissemination**

This study uses published aggregate data and does not require ethical approval. Findings will be disseminated in a peer-reviewed journal.

**Amendments**

In case of protocol amendments, the date of each amendment will be accompanied by a description of the change and the rationale.

**Abbreviations**

AMR: antimicrobial resistance

CAP: community-acquired pneumonia

DE-NMA: dose-effect network meta-analysis

ITT: intention-to-treat

PP: per protocol

PRISMA: Preferred Reporting Items for Systematic Reviews and Meta-analyses

**2. Search strings used for Ovid MEDLINE, Embase, and CENTRAL**

2-1. Search strategy for Ovid MEDLINE

1 randomized controlled trial.pt.

2 controlled clinical trial.pt.

3 randomized.ab.

4 placebo.ab.

5 drug therapy.fs.

6 randomly.ab.

7 trial.ab.

8 groups.ab.

9 or/1-8

10 exp animals/ not humans.sh.

11 9 not 10

12 exp Community-Acquired Infections/

13 Pneumonia, Bacterial/dt [Drug Therapy]

14 community acquired pneumonia.ab,ti.

15 (12 and 13) or 14

16 ((short adj term) or (long adj term) or prolonged or (short adj course) or (long adj course) or day or days or duration or disconti*).mp.

17 (beta-lactam* or macrolide* or quinolone* or tetracycline* or amikacin or amoxicillin or ampicillin or azithromycin or cefepim or cefotaxim* or ceftarolin or ceftazidim* or ceftibuten or ceftriaxon* or cefuroxim* or cethromycin or ciprofloxacin or clarithromycin or clavulanic acid or clindamycin or co-amoxiclav or co-trimoxacol or doxycyclin* or ertapenem or erythromycin or fluoroquinolon* or fluorchinolon* or gemifloxacin or gentamicin or imipenem or levofloxacin or linezolide or meropenem or moxifloxacin or penicillin* or piperacillin or roxithromycin or sultamicillin or tazobactam or telithromycin or tetracyclin* or ticarcillin or tobramycin).mp.

18 Anti-Bacterial Agents/ad [Administration & Dosage]

19 17 or 18

20 11 and 15 and 16 and 19

2-2. Search strategy for Embase

S1	(EMB.EXACT.EXPLODE("community acquired infection")) AND (EMB.EXACT("bacterial pneumonia -- drug therapy"))

S2	ab(community acquired pneumonia) OR ti(community acquired pneumonia)

S3	S2 OR S1

S4	ab((short near/1 term) OR (long near/1 term) OR prolonged OR (short near/1 course) OR (long near/1 course) OR day OR days OR duration or disconti*) OR ti((short near/1 term) OR (long near/1 term) OR prolonged OR (short near/1 course) OR (long near/1 course) OR day OR days OR duration or disconti*)

S5	ab(beta-lactam* OR macrolide* OR quinolone* OR tetracycline* OR amikacin OR amoxicillin OR ampicillin OR azithromycin OR cefepim OR cefotaxim* OR ceftarolin OR ceftazidim* OR ceftibuten OR ceftriaxon* OR cefuroxim* OR cethromycin OR ciprofloxacin OR clarithromycin OR clavulanic acid OR clindamycin OR co-amoxiclav OR co-trimoxacol OR doxycyclin* OR ertapenem OR erythromycin OR fluoroquinolon* OR fluorchinolon* OR gemifloxacin OR gentamicin OR imipenem OR levofloxacin OR linezolide OR meropenem OR moxifloxacin OR penicillin* OR piperacillin OR roxithromycin OR sultamicillin OR tazobactam OR telithromycin OR tetracyclin* OR ticarcillin OR tobramycin) OR ti(beta-lactam* OR macrolide* OR quinolone* OR tetracycline* OR amikacin OR amoxicillin OR ampicillin OR azithromycin OR cefepim OR cefotaxim* OR ceftarolin OR ceftazidim* OR ceftibuten OR ceftriaxon* OR cefuroxim* OR cethromycin OR ciprofloxacin OR clarithromycin OR clavulanic acid OR clindamycin OR co-amoxiclav OR co-trimoxacol OR doxycyclin* OR ertapenem OR erythromycin OR fluoroquinolon* OR fluorchinolon* OR gemifloxacin OR gentamicin OR imipenem OR levofloxacin OR linezolide OR meropenem OR moxifloxacin OR penicillin* OR piperacillin OR roxithromycin OR sultamicillin OR tazobactam OR telithromycin OR tetracyclin* OR ticarcillin OR tobramycin)

S6	(EMB.EXACT("antibiotic agent -- drug dose"))

S7	S6 OR S5

S8	S7 AND S4 AND S3

S9	(ab(random*) OR ti(random*)) OR (ab(placebo*) OR ti(placebo*)) OR (ab(double NEAR/1 blind*) OR ti(double NEAR/1 blind*))

S10	S9 AND S8

2-3. Search strategy for CENTRAL

#1	[mh "Community-Acquired Infections"]

#2	[mh "Pneumonia, Bacterial"]

#3	"community acquired pneumonia":ti,ab

#4	(#1 and #2) or #3

#5	(short:ti,ab,kw NEXT term:ti,ab,kw) OR (long:ti,ab,kw NEXT term:ti,ab,kw) OR prolonged:ti,ab,kw OR (short:ti,ab,kw NEXT course:ti,ab,kw) OR (long:ti,ab,kw NEXT course:ti,ab,kw) OR day:ti,ab,kw OR days:ti,ab,kw OR duration:ti,ab,kw OR disconti*:ti,ab,kw

#6	beta-lactam*:ti,ab,kw OR macrolide*:ti,ab,kw OR quinolone*:ti,ab,kw OR tetracycline*:ti,ab,kw OR amikacin:ti,ab,kw OR amoxicillin:ti,ab,kw OR ampicillin:ti,ab,kw OR azithromycin:ti,ab,kw OR cefepim:ti,ab,kw OR cefotaxim*:ti,ab,kw OR ceftarolin:ti,ab,kw OR ceftazidim*:ti,ab,kw OR ceftibuten:ti,ab,kw OR ceftriaxon*:ti,ab,kw OR cefuroxim*:ti,ab,kw OR cethromycin:ti,ab,kw OR ciprofloxacin:ti,ab,kw OR clarithromycin:ti,ab,kw OR "clavulanic acid":ti,ab,kw OR clindamycin:ti,ab,kw OR co-amoxiclav:ti,ab,kw OR co-trimoxacol:ti,ab,kw OR doxycyclin*:ti,ab,kw OR ertapenem:ti,ab,kw OR erythromycin:ti,ab,kw OR fluoroquinolon*:ti,ab,kw OR fluorchinolon*:ti,ab,kw OR gemifloxacin:ti,ab,kw OR gentamicin:ti,ab,kw OR imipenem:ti,ab,kw OR levofloxacin:ti,ab,kw OR linezolide:ti,ab,kw OR meropenem:ti,ab,kw OR moxifloxacin:ti,ab,kw OR penicillin*:ti,ab,kw OR piperacillin:ti,ab,kw OR roxithromycin:ti,ab,kw OR sultamicillin:ti,ab,kw OR tazobactam:ti,ab,kw OR telithromycin:ti,ab,kw OR tetracyclin*:ti,ab,kw OR ticarcillin:ti,ab,kw OR tobramycin:ti,ab,kw

#7	[mh "Anti-Bacterial Agents"]

#8	#6 OR #7

#9	#4 AND #5 AND #8

**3. Amendments from the protocol**

We reconsidered data structure and realized that dose-effect meta-analysis, not *network* meta-analysis would be more suitable. We also realized that the small number of included studies would make using four or more knots inappropriate and decided not to conduct sensitivity analyses with different number of knots. We searched Embase via ProQuest in addition to MEDLINE and CENTRAL. (25th August, 2021, before starting formal screening)

We additionally extracted baseline severity data using Pneumonia Severity Index (10th October, 2021, after full text screening done, before data extraction started).

We planned to conduct a sensitivity analysis excluding trials with inpatients, but we found only one trial focusing on outpatients. We therefore decided to conduct a sensitivity analysis excluding trials with outpatients instead. (25th October, 2021, after data extraction)

We additionally conducted a sensitivity analysis excluding trials which randomized patients after achieving clinical stability. (27th October, 2021, after data extraction. Post hoc)

**4. List of all included papers**

- Siegel RE, Alicea M, Lee A, Blaiklock R. Comparison of 7 Versus 10 Days of Antibiotic Therapy for Hospitalized Patients with Uncomplicated Community-Acquired Pneumonia. *Am J Ther* 1999; 6: 217–22.
- Léophonte P, Choutet P, Gaillat J, et al. Efficacité comparée de la ceftriaxone dans un traitement de dix jours versus un traitement raccourci de cinq jours des pneumonies aigues communautaires de l’adulte hospitalisé avec facteur de risque. *Médecine Et Maladies Infect* 2002; 32: 369–81.
- Tellier G, Niederman MS, Nusrat R, et al. Clinical and bacteriological efficacy and safety of 5 and 7 day regimens of telithromycin once daily compared with a 10 day regimen of clarithromycin twice daily in patients with mild to moderate community-acquired pneumonia. *J Antimicrob Chemoth* 2004; 54: 515–23.
- El Moussaoui R, Borgie C, Broek P, et al. Effectiveness of discontinuing antibiotic treatment after three days versus eight days in mild to moderate-severe community acquired pneumonia: randomised, double blind study. *BMJ* 2006; 332: 1355.
- File TM, Mandell LA, Tillotson G, et al. Gemifloxacin once daily for 5 days versus 7 days for the treatment of community-acquired pneumonia: a randomized, multicentre, double-blind study. *J Antimicrob Chemoth* 2007; 60: 112–20.
- Strålin K, Rubenson A, Lindroth H, et al. BETALACTAM TREATMENT UNTIL NO FEVER FOR 48HOURS (AT LEAST 5 DAYS) VERSUS 10 DAYS IN COMMUNITY-ACQUIRED PNEUMONIA: RANDOMISED, NON-INFERIORITY, OPEN STUDY. *Pneumonia* 2014; 3: 246–81.
- Uranga A, España PP, Bilbao A, et al. Duration of Antibiotic Treatment in Community-Acquired Pneumonia: A Multicenter Randomized Clinical Trial. *JAMA Intern Med* 2016; 176: 1257.
- Aliberti S, Ramirez J, Giuliani F, et al. Individualizing duration of antibiotic therapy in community-acquired pneumonia. *Pulm Pharmacol Ther* 2017; 45: 191–201.
- Dinh A, Ropers J, Duran C, et al. Discontinuing β-lactam treatment after 3 days for patients with community-acquired pneumonia in non-critical care wards (PTC): a double-blind, randomised, placebo-controlled, non-inferiority trial. *Lancet* 2021; 397: 1195–203.

**Ongoing trials**

- NCT03609099. Adequate Duration of Antibiotic Treatment in Community-acquired Pneumonia With High Risk Class and Adequate Initial Clinical Response (2017-001406-15). Available from: https://clinicaltrials.gov/ct2/show/NCT03609099
- NCT04089787. Shortened Antibiotic Treatment of 5 Days in Community-Acquired Pneumonia (CAP5). Available from: https://clinicaltrials.gov/ct2/show/NCT04089787

**5. List of excluded studies**

| **Name** | **Title** | **Comment** |
| --- | --- | --- |
| EUCTR2005-000105-65 | Comparative study of the efficacy and tolerance of intravenously administered azithromycin (1.5 g) given either as a single dose or over a 3 day period in patients with community-acquired pneumonia | wrong intervention (dfferent drugs) |
| EUCTR2014-003137-25 | Optimal duration of antibiotic treatment in patients with complicated parapneumonic pleural effusions or empyema | wrong intervention (dfferent drugs) |
| EUCTR2020-004452-15 | ADMINISTRATION OF CLARITHROMYCIN IN COMMUNITY-ACQUIRED PNEUMONIA | wrong intervention (dfferent drugs) |
| Fekete2021 | In moderately severe CAP stable after 3 d of beta-lactam, stopping therapy was noninferior to 5 additional d. | wrong design (comment) |
| File2007 | No Title (Author's reply) | wrong design |
| Fine2003 | Implementation of an evidence-based guideline to reduce duration of intravenous antibiotic therapy and length of stay for patients hospitalized with community-acquired pneumonia: a randomized controlled trial | wrong intervention (dfferent drugs) |
| JPRN-JapicCTI-163439 | A Phase III study of Solithromycin in patients with community-acquired pneumonia | wrong intervention (dfferent drugs) |
| JPRN-UMIN000008677 | Efficacy and Safety of treatment with Levofloxacin for Community-acquired Pneumonia | wrong design (single arm) |
| JPRN-UMIN000011835 | Efficacy and safety of meropenem (3g/day) in the treatment of severe/refractory respiratory infections | wrong design (single arm) |
| JPRN-UMIN000011836 | Efficacy and safety of azithromycin infusion in the treatment of mild/moderate community-acquired pneumonia | wrong design (observational) |

| **Name** | **Title** | **Comment** |
| --- | --- | --- |
| Li2007 | Efficacy of Short-Course Antibiotic Regimens for Community-Acquired Pneumonia: A Meta-analysis | wrong design (review) |
| Li2021 | A multicenter randomized controlled study on the efficacy of moxifloxacin and garenoxacin for the treatment of adult community-acquired pneumonia | wrong intervention (dfferent drugs) |
| Lyttle2019 | Dose and duration of antibiotic treatment in young children with community-acquired pneumonia | wrong participants |
| Malhotra-Kumar2016 | Impact of amoxicillin therapy on resistance selection in patients with community-acquired lower respiratory tract infections: a randomized, placebo-controlled study | wrong participants |
| Melo2018 | Shortening antibiotic duration for community acquired pneumonia. | wrong design (review) |
| Scalera2007 | How long should we treat community-acquired pneumonia?. | wrong design (review) |
| Stralin2004 | Short-course beta-lactam treatment for community-acquired pneumonia. | wrong design (review) |
| Uranga2015 | Duration of Antibiotic Treatment in Community-Acquired Pneumonia. | wrong design (review) |
| Vetter2002 | A prospective, randomized, double-blind multicenter comparison of parenteral ertapenem and ceftriaxone for the treatment of hospitalized adults with community-acquired pneumonia | wrong intervention (dfferent drugs) |
| Weber1987 | Ampicillin versus cefamandole as initial therapy for community-acquired pneumonia | wrong intervention (dfferent drugs) |
| YangJ2020 | The combined treatment of imipenem cilastatin and azithromycin for elderly patients with community-acquired pneumonia | wrong intervention (dfferent drugs) |

**6. Heterogeneity: Variance partition coefficient for the primary outcome**

VPC is computed for each non-referent arm of each study (those that have OR≠1). We included nine two-armed trials, and thus we have 9 VPC numbers. We present them below. It is generally interpreted as: VPC values below 25% low, 25-75% moderate and over 75% high.

```
>   vpc(mod1)
           2            4            6            8           10           12           14           16           18
1.059171e-10 1.102071e-09 3.592398e-09 4.059647e-09 2.000592e-09 8.322319e-10 1.771638e-09 1.071397e-10 1.843283e-08
```

**7. Sensitivity analyses**

Duration-effect relationship of secondary outcomes could not be computed due to missing data in some cases.

\# A priori sensitivity analyses

##S1 To test the stability of the shape of the spline curves, we used different locations of knots (10%, 50%, 90%).

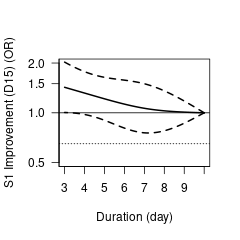

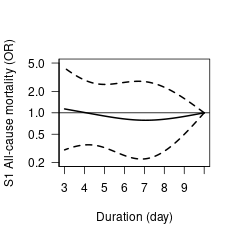

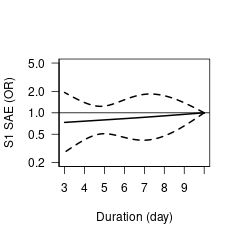

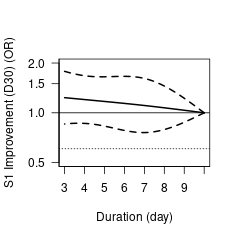

##S2.1 To test the influence of trials included, we conducted sensitivity analyses excluding trials with overall high risk of bias (excluding Siegel1999, Leophonte2002, Stralin2014, Aliberti2017)

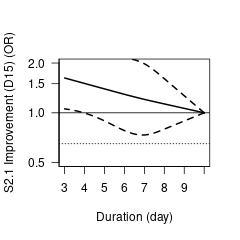

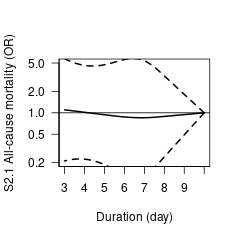

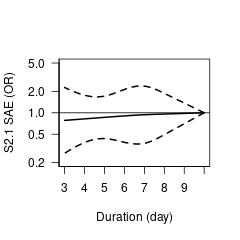

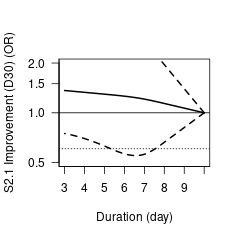

##S2.2 To test the influence of trials included, we conducted sensitivity analyses excluding trials with outpatients (excluding Tellier2004, File2007. SAE not computable)

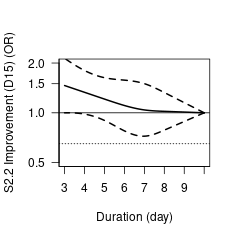

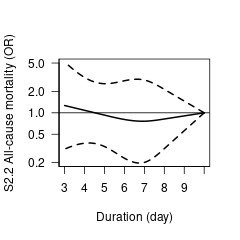

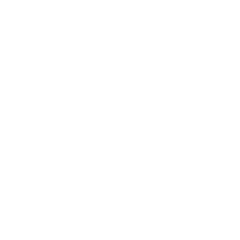

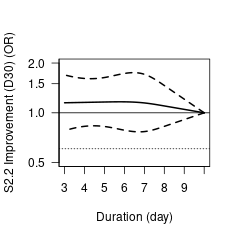

##S3 To test the robustness of the analytical method, we used PP dataset. (All-cause mortality and SAE not computable)
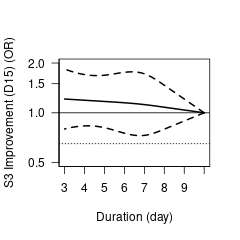

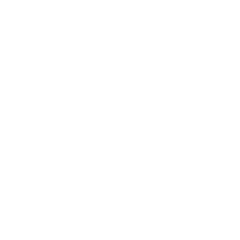

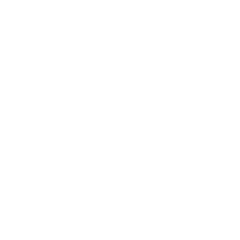

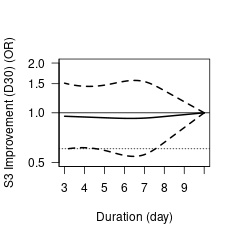

##S4 To test the influence of antibiotics examined, we conducted sensitivity analyses including only antibiotics recommended for empirical treatment of CAP by clinical guidelines. (excluding Siegel1999, Tellier2004. SAE not computable.We included trials that used various antibiotics)

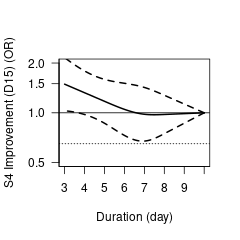

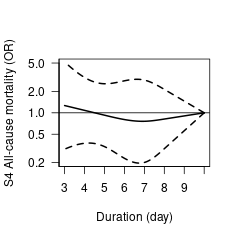

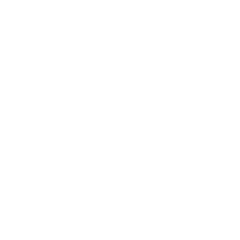

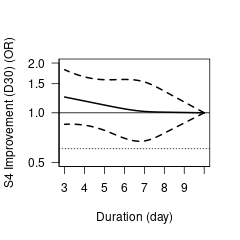

\# Post-hoc, exploratory sensitivity analyses

##S5.1 Randomization before the initial antibiotic treatment (including Siegel1999, Leophonete2002, Tellier2004, File2007, Stralin2014. SAE not computable)

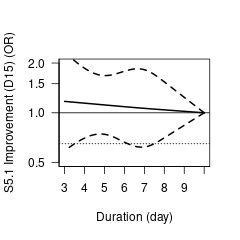

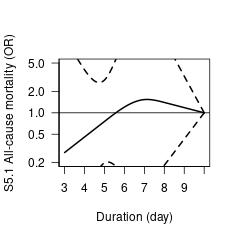

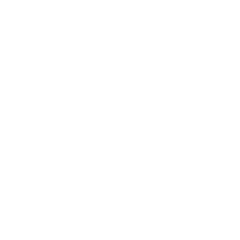

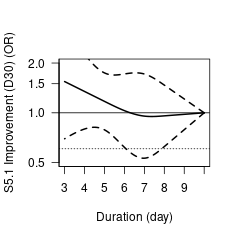

##S5.2 Randomization after several days or clinical stability achieved (including ElMoussaoui2006, Uranga2016, Aliberti2017, Dinh2021. SAE not computable)

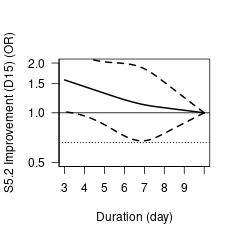

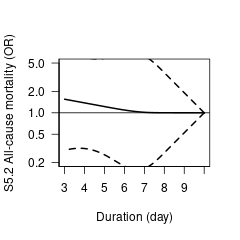

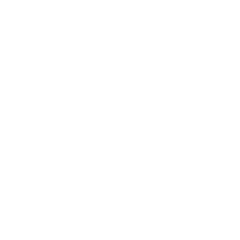

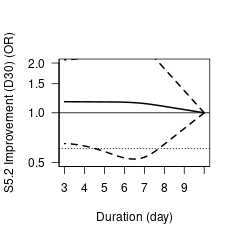
